# The relationship between mental health, sleep quality, and the immunogenicity of COVID-19 vaccinations

**DOI:** 10.1101/2023.02.26.23286402

**Authors:** Isabell Wagenhäuser, Julia Reusch, Alexander Gabel, Juliane Mees, Helmut Nyawale, Anna Frey, Thiên-Trí Lâm, Alexandra Schubert-Unkmeir, Lars Dölken, Oliver Kurzai, Stefan Frantz, Nils Petri, Manuel Krone, Lukas B. Krone

## Abstract

Sleep modulates the immune response and sleep loss can reduce the immunogenicity of certain vaccinations. Vice versa immune responses impact sleep. We aimed to investigate the influence of mental health and sleep quality on the immunogenicity of COVID-19 vaccinations and, conversely, of COVID-19 vaccinations on sleep quality.

The prospective CoVacSer study monitored mental health, sleep quality, and Anti-SARS-CoV-2-Spike IgG titres in a cohort of 1,082 healthcare workers from the 29^th^ of September 2021 to the 19^th^ of December 2022. Questionnaires and blood samples were collected before, 14 days, and three months after the third COVID-19 vaccination. In 154 participants the assessments were also conducted before and 14 days after the fourth COVID-19 vaccination.

Healthcare workers with psychiatric disorders had slightly lower Anti-SARS-CoV-2-Spike IgG levels before the third COVID-19 vaccination. However, this effect was mediated by higher median age and body mass index in this subgroup. Antibody titres following the third and fourth COVID-19 vaccination (‘booster vaccinations’) were not significantly different between subgroups with and without psychiatric disorders. Sleep quality did not affect the humoral immunogenicity of the COVID-19 vaccinations. Moreover, the COVID-19 vaccinations did not impact self-reported sleep quality.

Our data suggests that in a working population neither mental health nor sleep quality relevantly impact the immunogenicity of COVID-19 vaccinations and that COVID-19 vaccinations are not a precipitating factor for insomnia. The findings from this large-scale real-life cohort study will inform clinical practice regarding the recommendation of COVID-19 booster vaccination for individuals with mental health and sleep problems.

## 1. Introduction

The immunogenicity of vaccinations is modulated by a plethora of physiological and behavioural influences (Zimmermann & Curtis, 2019). A bidirectional relationship between sleep and immune function is well established (Besedovsky et al., 2019). Sleep can modulate the cellular and humoral response to vaccinations (Lange et al. 2011). Vice versa, vaccinations can impact sleep (Sharpley et al., 2016) and immune system activation might be involved in the pathophysiological hyperarousal of insomnia patients (Riemann et al., 2020; Riemann et al., 2010).

Previous work on the impact of sleep on immunogenicity of different vaccinations has found that experimental sleep restriction and sleep deprivation as well as habitual short sleep impair the humoral immune response following influenza, hepatitis A, and hepatitis B vaccinations (Rayatdoost et al., 2022). The most extensive study monitored antibody levels for one year over a course of three vaccinations against hepatitis A and showed that whole night of sleep deprivation after each of the vaccinations significantly reduced antibody levels in the short and long term (Lange et al., 2011), supporting previous findings of reduced hepatitis A antibody levels four weeks after a single hepatitis A vaccination was followed by experimental sleep deprivation (Lange et al., 2003). However, for other vaccines the effects of sleep deprivation are less clear. Volunteers undergoing six days of bedtime restriction around a seasonal influenza vaccination had only half of the antibody levels at 10 days post vaccination compared to normally sleeping volunteers, but 3-4 weeks following the sleep restriction antibody levels no longer differed between groups (Spiegel et al., 2002). Similarly, one night of sleep deprivation following an influenza H1N1 vaccination impacted the antibody levels only in the five-day follow up in males and while there was no difference to controls in females or males at later time points (Benedict et al., 2012). The relationship between short sleep and reduced vaccination response was corroborated by two studies investigating the effect of sleep habits immunogenicity of hepatitis B (Prather et al., 2012) and a tetravalent influenza vaccination (Prather et al., 2021). However, in both studies neither sleep efficiency nor sleep quality mediated antibody levels. Similarly, sleep disruptions due to obstructive sleep apnoea did not affect antibody levels (Dopp et al., 2007). If sleep disorders impact vaccination immunogenicity also remains unclear following a study on influenza vaccination in insomnia patients (Taylor et al., 2017). This study found lower antibody levels in insomnia patients at baseline and after the vaccination as well as some indications from exploratory analyses that insomnia and the Pittsburgh Sleep Quality Index (PSQI) might mediate the vaccination response (Buysse et al., 1989). However, the main analysis showed no interaction effect between the time point (before vs. after vaccination) and the group (insomnia vs. controls).

The COVID-19 vaccination has become a key prevention tool in the ongoing COVID-19 pandemic (Benenson et al., 2021). It has been speculated that poor sleep quality may impair the COVID-19 vaccination derived humoral immune response (Kow & Hasan, 2021; Zhu et al., 2021), particularly in individuals with psychiatric disorders (Mazereel et al., 2021). However, data on the relationship between sleep and immunogenicity of COVID-19 vaccinations is still sparse. To our knowledge, currently the first has recently been conducted in a small cohort of Greek healthcare workers (HCWs) (Athanasiou et al., 2023). Surprisingly, night shifts two days before or one day after COVID-19 vaccination did not affect antibody levels. However, the Athens Insomnia Scale (AIS) (Soldatos et al., 2000) and the PSQI predicted antibody levels after the first and second vaccination (Athanasiou et al., 2023), yet to a smaller degree than other established factors such as age and smoking. Considering that the study participants were assessed during the peak of the COVID-19 pandemic and severely sleep deprived with an average sleep duration of only six hours per night, it remains unclear if the effects were mediated by sleep duration, sleep quality or other factors such as mental health.

Mental health, however, is another important aspect to consider in the vaccination response (Mazereel et al., 2021). Some mental health conditions, in particular major depressive disorder, can reduce vaccine efficacy (Xiao et al., 2022). Yet, the current evidence regarding the effect of psychiatric disorders on vaccine immunogenicity is heterogenous and data on the COVID-19 vaccination is missing (Mazereel et al., 2021; Xiao et al., 2022). Considering that the COVID-19 pandemic has now been ongoing for more than three years (Wu et al., 2020), and the novel mRNA-based vaccines (Baden et al., 2020; Polack et al., 2020) have been administered to billions of individuals worldwide, it is surprising that the impact of sleep and mental health on the COVID-19 vaccine immunogenicity and the impact of the vaccination on sleep have not yet been studied in a large cohort.

We set out to use our large-scale real-life data of 1,082 HCWs undergoing their third and fourth COVID-19 vaccination (first and second booster vaccination) to address this question. The sample of HCWs is particularly relevant for vaccine recommendations as this population is, on one hand, highly exposed to SARS-CoV-2 and needs the best possible vaccination protection (Gross et al., 2021; Montgomery et al., 2021), but on the other hand, predisposed to poor sleep and mental stress due to their daily work (Anderson et al., 2012; Chang et al., 2013; Wolkow et al., 2015). We therefore tested in this sample if mental health and sleep quality affect the immune response following COVID-19 vaccination and, vice versa, whether the COVID-19 vaccination affects sleep quality.

## 2. Methods

### 2.1 Study setting

This project was part of the CoVacSer cohort study which investigates prospectively the SARS-CoV-2 immunity, quality of life, and ability to work in HCWs after COVID-19 vaccination and/or SARS-CoV-2 infection. Inclusion criteria for study enrolment were: (i) age ≥ 18 years, (ii) written consent form, (iii) 14 days minimum interval after the first polymerase chain reaction (PCR) confirmed SARS-CoV-2 infection and/or at least one dose of COVID-19 vaccination independent of vaccination regime, (iv) employment in healthcare sector. Individuals that received vaccines without European Medicines Agency (EMA) authorisation during the data collection period were excluded from the data analysis according to the study protocol.(European Medicines Agency (EMA), 2023)

### 2.2 Data collection

The data was collected between the 29^th^ of September 2021 to the 19^th^ of December 2022. Analysis was performed based on the vaccination status:

A. third mRNA-based COVID-19 vaccination (first booster dose)
B. fourth mRNA-based COVID-19 vaccination (second booster dose)

The following COVID-19 vaccines were used:

i. monovalent BNT162b2mRNA (Comirnaty, BioNTech/Pfizer, Mainz/Germany, New York/USA; full dose 30µg mRNA)
ii. mRNA-1273 (Spikevax, Moderna, Cambridge/USA; half dose 50g mRNA as booster dose following the recommendations of the German constant vaccination committee (STIKO) (Robert Koch-Institut (RKI), 2021)
iii. bivalent BNT162b2 mRNA Original/Omicron BA.1 (Comirnaty Original/Omicron BA.1, BioNTech/Pfizer, Mainz/Germany, New York/USA)
iv. bivalent BNT162b2 mRNA Original/Omicron BA.4-5 (Comirnaty Original/Omicron BA.4-5, BioNTech/Pfizer, Mainz/Germany, New York/USA)

The study participants submitted the CoVacSer study survey with a corresponding serum blood sample at each participation date, which were defined in the study protocol:

1. **pre-vaccination** baseline assessment before the respective COVID-19 booster vaccine administration
2. assessment 14 to 30 days after each booster vaccination, referred to as **14-days post-vaccination participation**.
3. follow-up assessment 60 to 120 days after booster vaccination, referred to as **3**-**month follow**-**up**.

Because most participants who received a fourth vaccination dose were vaccinated less than three months before the end date of the data collection period, the 3-month follow-up data was only analysed for the third COVID-19 vaccination.

The CoVacSer study survey includes socio-demographic aspects and individual risk factors, containing the World Health Organisation Quality of Life (WHOQOL-BREF) questionnaire as well as the Work Ability Index (WAI) (Gholami et al., 2013; Skevington, 1999; van den Berg et al., 2009).

The ability to enjoy life, the ability to concentrate, energy for everyday life, and acceptance of one’s own appearance were asked in the categories “not at all”, “a little”, “average”, “quite” and “extremely”. Satisfaction with the ability to cope with everyday life, self-satisfaction, satisfaction with sexual life, as well as the quality of sleep were queried in the categories “very dissatisfied”, “dissatisfied”, “neither satisfied nor dissatisfied”, “satisfied”, and “very satisfied”. The frequency of occurrence of negative feelings in the frequency intervals “never”, “not often”, “occasionally”, “often”, and “always”. Participants werer instructed to rate all items for 14-days period prior to the respective assessment date.

For the analysis on the impact of mental health on vaccine immunogenicity, study participants without psychiatric disorders were compared to those who indicated to have a psychiatric disorder. To determine whether or not participants had a psychiatric disorder, the questionnaire entry 14 days after the third or fourth vaccination was used. For the analysis of the effect of sleep quality on the humoral immunogenicity of the COVID-19 vaccination, the sleep quality reported at the 14-days assessment after vaccination was used, as this refers to the time span immediately following the vaccination.

Only blood samples with a signed written consent form and a completed linked questionnaire were considered. REDCap (Research Electronic Data Capture, projectredcap.org) was used as technological platform for the questionnaire recording (Harris et al., 2019; Harris et al., 2009). In the context of pseudonymisation, blood samples were assigned to the study survey based on date of birth and dates of COVID-19 vaccination.

Most HCWs were recruited from a single tertiary hospital in Germany, the University Hospital Würzburg, along with some HCWs from surrounding hospitals and medical surgeries.

### 2.3 SARS-CoV-2 IgG ELISA

The measurement of Anti-SARS-CoV-2-Spike IgG levels was performed using the SERION ELISA *agile* SARS-CoV-2 IgG (SERION diagnostics, Würzburg, Germany) as an enzyme linked immunoassay. Detected extinction values were converted to Serion IgG units per ml (U/ml) as producer specific units in use of the software easyANALYSE (SERION diagnostics). Consequently, the internationally established unit Binding Antibody Units per ml (BAU/ml) were calculated using the factor 2.1 in accordance with manufacturer’s information. For the measurement of Anti-SARS-CoV-2-Spike-IgG levels beyond the maximum limit of 250 U/ml (>525 BAU/mL), a dilution series containing dilution factors both 10 and 100 was conducted consistent with previous studies (Krone et al., 2021; Perkmann et al., 2021).

### 2.4 Ethical approval

The study protocol was approved by the Ethics committee of the University of Würzburg in accordance with the Declaration of Helsinki (file no. 79/21).

### 2.5 Statistics

Data analysis was performed using GraphPad Prism 9.5.1 (GraphPad Software, San Diego, CA, USA). For categorical variables the statistical significance levels were calculated using Fisher’s exact tests (gender composition of the study cohort in total and separated by mental health status). For Anti-SARS-CoV-2-Spike IgG levels separated by psychiatric disorder status the statistical significance levels were calculated using the Mann-Whitney U test. The correlation of sleep quality and Anti-SARS-CoV-2-IgG levels was performed using a Spearman-Rank correlation. A multiple regression model was used to evaluate the effect of gender, age, BMI, smoking, SARS-CoV-2 convalescence, number of individuals living within the same household, and days since the last COVID-19 immunising event at the baseline study participation on Anti-Spike-SARS-CoV-2 IgG levels. The two-tailed significance level α was set to 0.05.

## 3. Results

### 3.1 Participant recruitment and characterisation of the study population

From 29^th^ of September 2021 to the 19^th^ of December 2022, 1,158 CoVacSer study participants submitted a serum blood sample along with the study questionnaire 14 to 30 days after their third COVID-19 vaccination. All individuals met the inclusion criteria. 76 participants were excluded because they had not participated in the study prior to the third vaccination, so the third COVID-19 vaccination cohort consisted of 1,082 HCWs. Of the 155 HCWs who participated 14 to 30 days after the fourth COVID-19 vaccination, one HCW was excluded due to missing baseline participation prior to the fourth vaccination. 154 HCWs could be eventually included for the cohort of the fourth COVID-19 vaccination (*Figure 1*).

**Figure 1:**
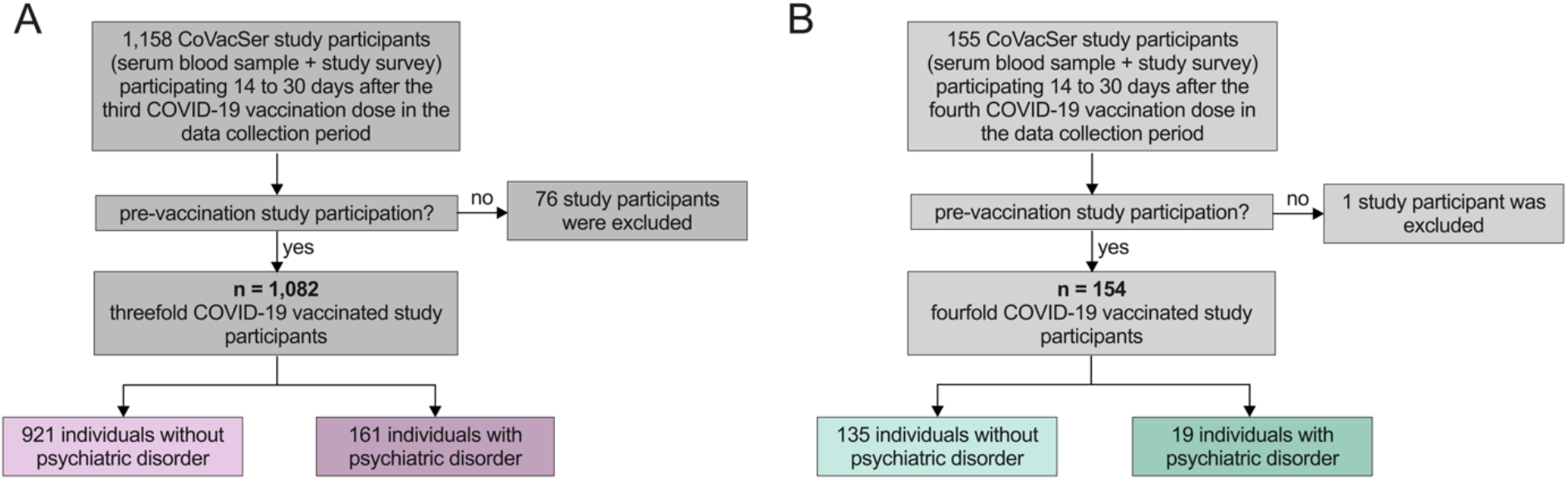
Recruitment of study participants A) Third COVID-19 vaccination. B) Fourth COVID-19 vaccination

For the third and fourth vaccination, assessments before and 14 days after the COVID-19 vaccination were conducted in all participants that received the respective vaccination. The 3-month follow-up after the third vaccination could be conducted in 785 (72.6%) individuals.

In the cohort of the **third COVID-19** vaccination (n=1,082 individuals), 86.0% (921) participants reported no, 14.0% (161) reported at least one psychiatric disorder. Separated by underlying psychiatric disorder, there was no significant difference in terms of gender composition, yet a clear trend towards a higher prevalence of psychiatric disorders in women (p=0.051, Fisher’s exact test, *Table 1*). HCWs with psychiatric disorders were on average older and had a higher body weight (both p<0.001, Mann-Whitney U test, *Table 1*) compared to those without psychiatric disorders, in line with the high comorbidity rates between obesity and mental health disorders (Avila et al., 2015). They also lived with fewer people in the same household (p=0.02, Mann-Whitney U test. We found no significant difference in the proportion of smokers among individuals with or without psychiatric disorders (p=0.40, Mann-Whitney U test; *Table 1*). In addition, the questionnaire assessment showed that participants with psychiatric disorders are less able to enjoy their lives, less able to concentrate, have less energy for daily life, lower self-acceptance of their appearance, an impaired ability to cope with everyday life, less self-satisfaction, lower satisfaction with their sexual life, and have more often negative feelings (all p<0.001, Mann-Whitney U test, Supplementary table 1).

**Table 1:** Characterisation of the subgroups without and with psychiatric disorders undergoing a third COVID-19 vaccination. Age, BMI, the interval since the last COVID-19 vaccination, intermission between the pre- and the initial post-vaccination participation as well as the number of household members are reported as medians with interquartile ranges in brackets. For each sub-cohort absolute numbers in the left, relative share in the right column. BMI: body mass index.

| study participants threefold COVID-19 vaccinated |  |  |  |  |  |  |  |  |
| --- | --- | --- | --- | --- | --- | --- | --- | --- |
|  |  | mentally healthy<br>(n=921) |  | underlying<br>psychiatric disorder<br>(n=161) |  | total (N=1,082) |  | p |
|  |  | absolute | relative | absolute | relative | absolute<br>number | relative<br>number |  |
| gender | female | 761 | 82.6% | 143 | 88.8% | 904 | 83.5% | 0.051 |
|  | male | 160 | 17.4% | 18 | 11.2% | 178 | 16.5% |  |
| age [years] | | 41 (31-52) | | 49 (37-58) | | 42 (31-54) | | $p<0.001$ |
| BMI [kg/m<sup>2</sup>] | | 23.7<br>(21.5-26.6) | | 25.5<br>(22.0-30.8) | | 23.8<br>(21.6-27.2) | | $p<0.001$ |
| smoking individuals |  | 323 | 35.1% | 62 | 38.5% | 385 | 35.6% | 0.40 |
| first post-vaccination<br>participation: interval<br>since third COVID-19<br>vaccination [days] |  | 17 (15-20) |  | 17 (15-21) |  | 17 (15-20) |  | 0.47 |
| interval of the prior<br>COVID-19 immunising<br>event to the baseline<br>participation [days] |  | 236<br>(203-266) |  | 236 (207-261) |  | 236 (203-265) |  | 0.75 |
| number of SARS-CoV-2<br>convalescent individuals |  | 30 | 3.3.% | 14 | 8.7% | 44 | 4.1% | 0.0013 |
| household members<br>[absolute number] |  | 2 (2-3) |  | 3 (1-3) |  | 2 (2-3) |  | 0.02 |

In the cohort of the **fourth COVID-19 vaccination** (n=154 individuals), 87.7% (135) participants reported no psychiatric disorder, 14.1% (19) HCWs reported at least one. In this much smaller cohort, we found no significant difference in any of the demographic items. However, again, individuals with psychiatric disorders tended to have a higher BMI (p=0.06, Mann-Whitney U-test, *Table 2*). Consistent with their mental health status, participants with psychiatric disorders indicated that they are less able to enjoy their lives, less able to concentrate, have less energy for daily life, lower self-acceptance of their appearance, an impaired ability to cope with everyday life, less self-satisfaction, and have more often negative feelings (all p<0.001, Mann-Whitney U test, *Supplementary Table 2*) as well as reduced satisfaction with their sexual life (p=0.0149, Mann-Whitney U test; Supplementary *Table 2*).

**Table 2:**
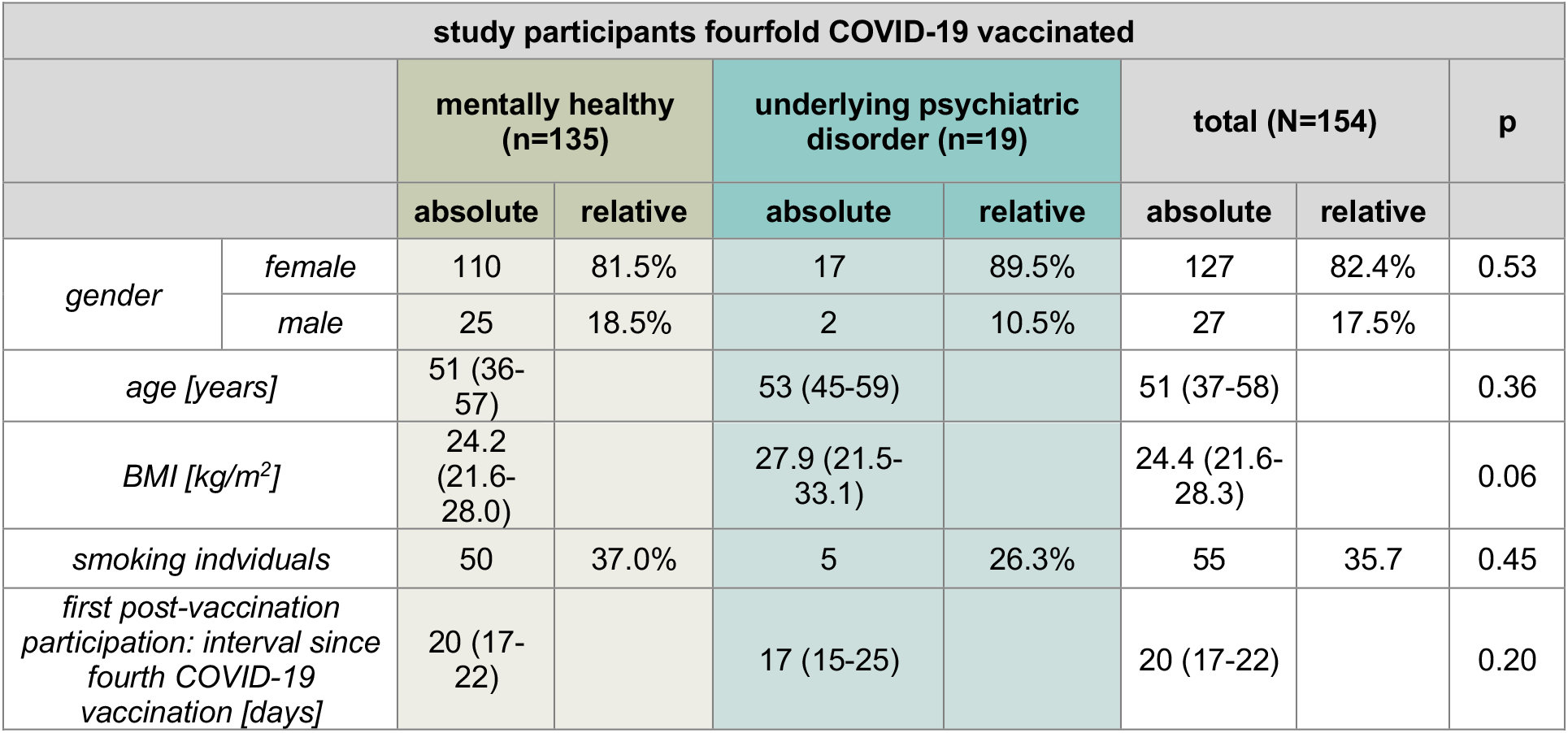
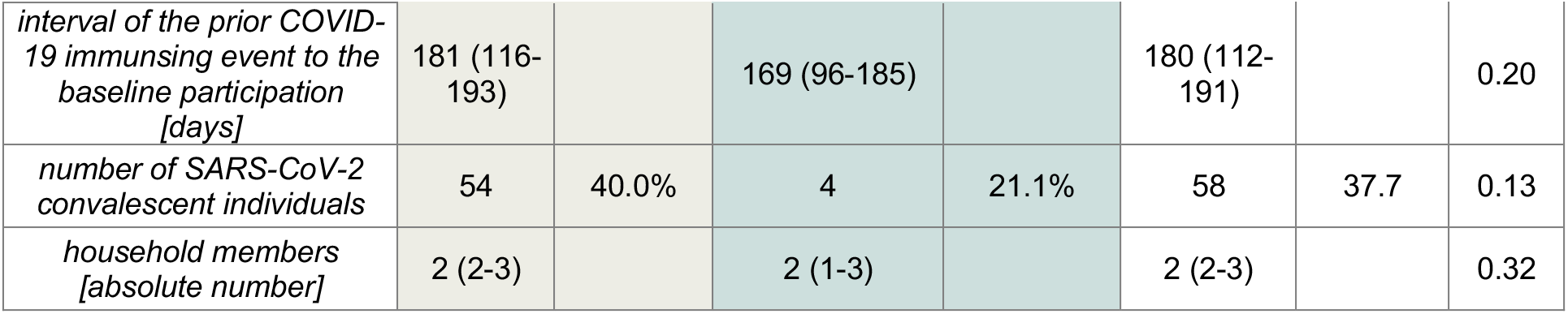
Characterisation of the subgroups without and with psychiatric disorders undergoing a fourth COVID-19 vaccination Age, BMI, the interval since the last COVID-19 vaccination, intermission between the pre- and the initial post-vaccination participation as well as the number of household members are reported as medians with interquartile ranges in brackets. For each sub-cohort absolute numbers in the left, relative share in the right column. BMI: body mass index.

### 3.2 Influence of mental health on post-vaccination Anti-SARS-CoV-2 spike IgG levels

To address the question of whether mental health impacts the Anti-SARS-CoV-2 spike IgG levels following COVID-19 vaccinations, we compared those study participants that reported no psychiatric disorder with those that indicated a psychiatric disorder.

In the **third COVID-19 vaccination** cohort, the median Anti-SARS-CoV-2-Spike IgG levels at baseline before the third vaccination were 149 (IQR:80 -259) BAU/ml for participants without and 119 (IQR:70-228) BAU/ml for participants with a psychiatric disorder. Hence, at baseline before the third vaccination, Anti-SARS-CoV-2-Spike IgG levels were significantly lower among HCWs with underlying psychiatric disorder (p=0.02, Mann-Whitney U test), while there was no significant difference in Anti-SARS-CoV-2-Spike IgG levels at the time points 14-days (p=0.20, Mann-Whitney U test) and 3-month (p=0.56, Mann-Whitney U test) after the third COVID-19 vaccination (*Figure 2A)*. To investigate potential mediators for the significant difference in Anti-SARS-CoV-2-Spike IgG levels between participants without and with psychiatric disorders at baseline before the third COVID-19 vaccination, we performed an explorative analysis using a multiple linear regression analysis. Age (p=0.03), SARS-CoV-2 infection convalescence (p<0.0001), and the interval between the last COVID-19 immunising event (defined as latest administration of a COVID-19 vaccine or SARS-CoV-2 infection; p<0.0001) significantly affected the Anti-SARS-CoV-2-Spike IgG levels, with higher age, absence of previous infection, and longer intervals to the last immunising event predicting lower antibody titres. Importantly, an underlying psychiatric disorder did not significantly influence the humoral immune response (p=0.69).

**Figure 2:**
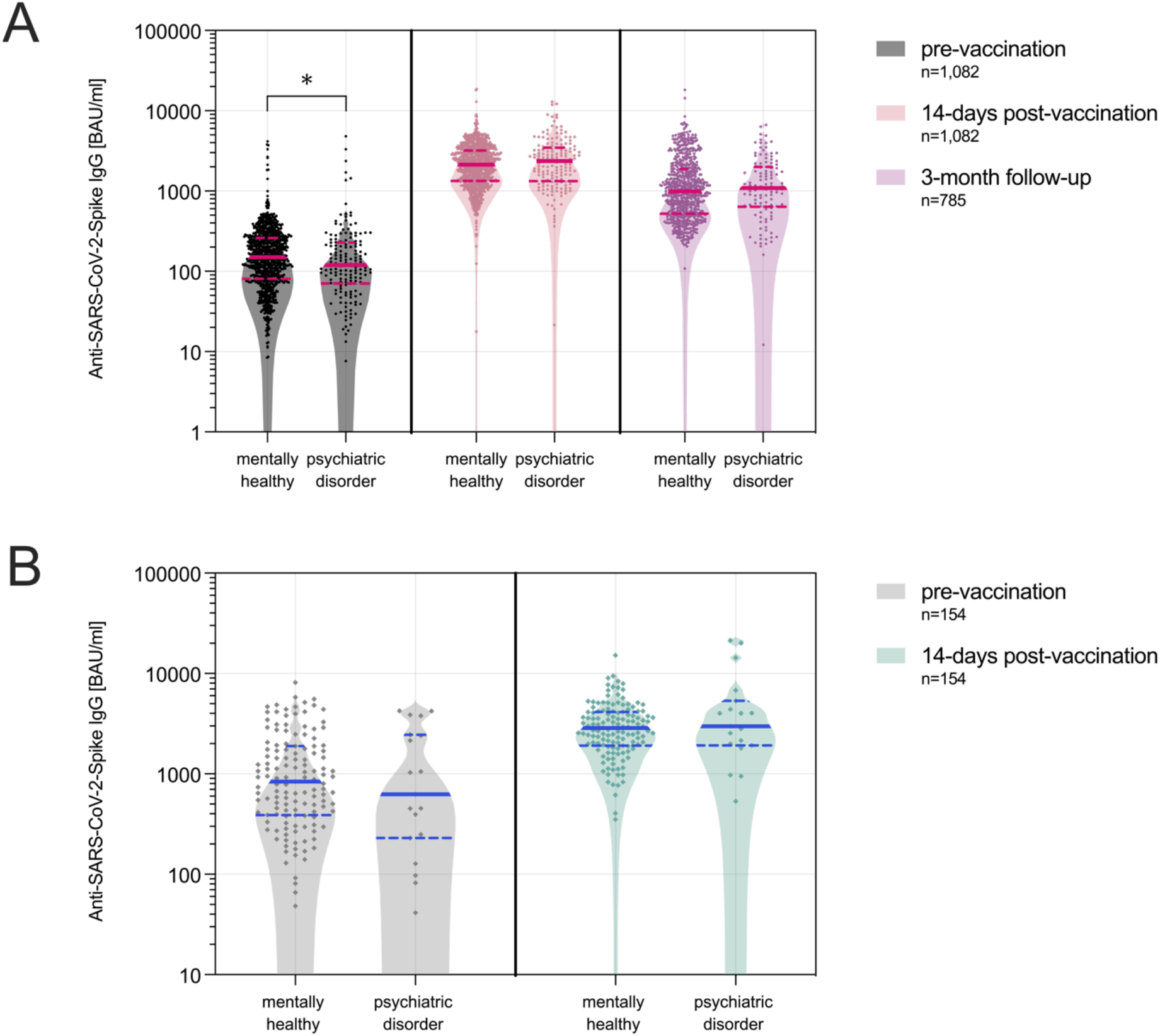
Anti-SARS-CoV-2-Spike IgG levels following COVID–19 booster vaccinations stratified by mental health status. **A**) Anti-SARS-CoV-2-Spike IgG levels among HCWs before (pre-vaccination) and after (14-days and 3-month follow up) the third COVID-19 vaccination. **B**) Anti-SARS-CoV-2-Spike IgG levels among HCWs before (pre-vaccination) and after (14-days) the fourth COVID-19 vaccination. Anti-SARS-CoV-2-Spike IgG logarithmically scaled. BAU/ml: binding antibody units per millilitre. Medians are indicated as horizontal bold line, quartiles as dotted lines. *: p < 0.05

In the **fourth COVID-19 vaccination** cohort, there was no significant difference in antibody titres at baseline (p=0.59, Mann-Whitney U test) or 14 days after vaccination (p=0.48, Mann-Whitney U test) between participants without and with psychiatric disorders (*Figure 2B)*.

### 3.3 Influence of sleep quality on post-vaccination Anti-SARS-CoV-2 spike IgG levels

We investigated the effect of sleep quality on Anti-SARS-CoV-2 spike IgG levels by comparing the antibody levels stratified by self-reported sleep quality.

In the **third vaccination** cohort, sleep quality and Anti-SARS-CoV-2-Spike IgG levels neither correlated pre-vaccination (p=0.93, r=0.002, Spearman correlation), 14-days (p=0.23, r=0.04, Spearman correlation) nor 3-month post-vaccination (p=0.73, r=-0.012, Spearman correlation, *Figure 3A*). In **the fourth COVID-19 vaccination** cohort, there was also no correlation between sleep and Anti-SARS-CoV-2-Spike IgG levels pre-vaccination (p=0.11, r=-0.13, Spearman correlation) nor 14-days post-vaccination (p=0.44, r=-0.06, Spearman correlation, *Figure 3B*).

**Figure 3:**
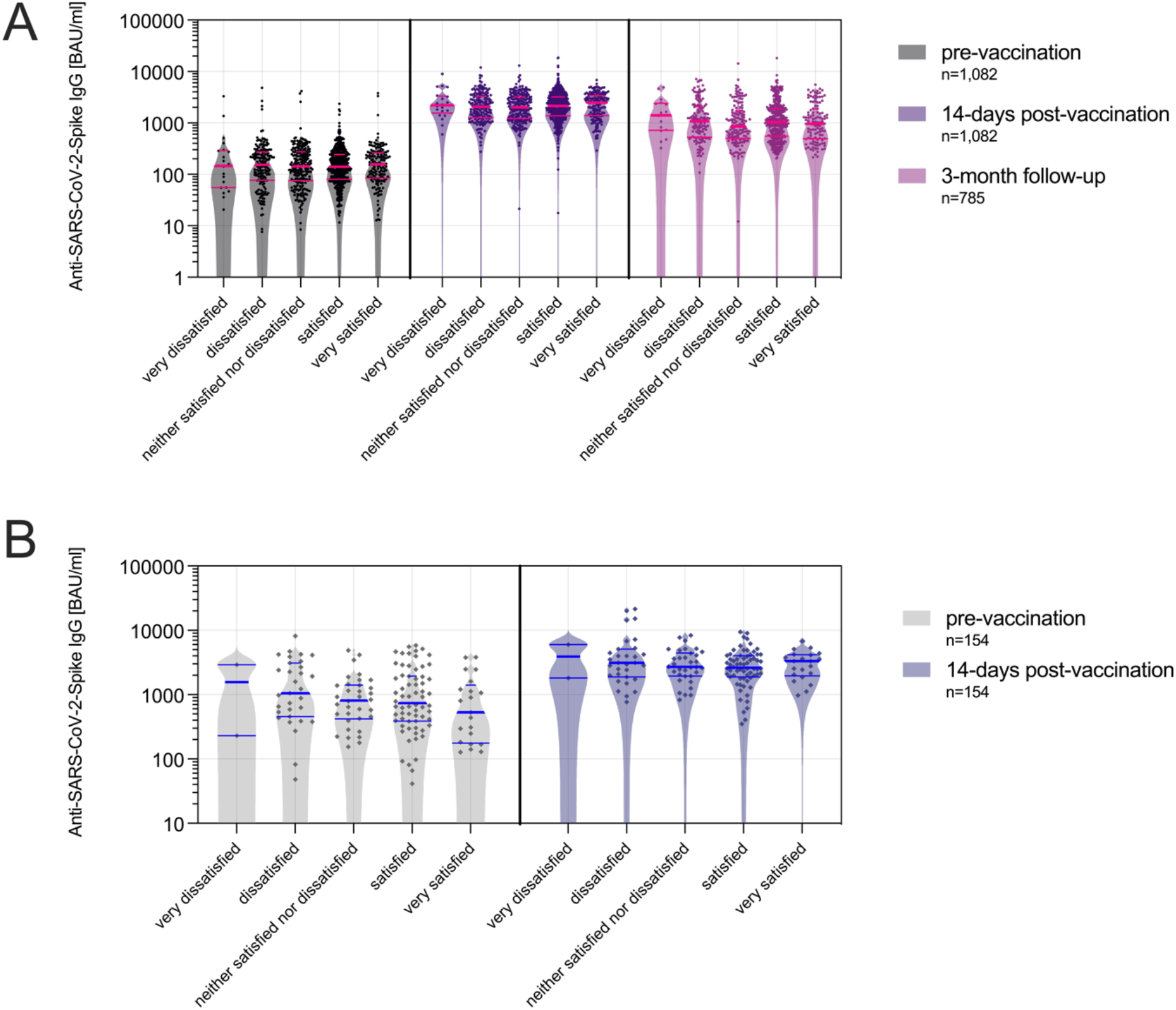
Anti-SARS-CoV-2-Spike IgG levels following COVID–19 booster vaccinations stratified by sleep quality. **A**) Anti-SARS-CoV-2-Spike IgG levels among HCWs before (pre-vaccination) and after (14-days and 3-month follow up) the third COVID-19 vaccination. **B**) Anti-SARS-CoV-2-Spike IgG levels among HCWs before (pre-vaccination) and after (14-days) the fourth COVID-19 vaccination. Sleep quality is stratified in five categories (very dissatisfied to very satisfied). Anti-SARS-CoV-2-Spike IgG logarithmically scaled. BAU/ml: binding antibody units per millilitre. Medians are indicated as horizontal bold line, quartiles as dotted lines.

### 3.4 Effect of COVID-19 vaccination on sleep quality

In order to assess whether the COVID-19 booster vaccinations affected sleep quality, we investigated the change of sleep quality across the time points before and after the third and fourth COVID-19 vaccination.

In the **third vaccination** cohort, there was no significant change in sleep quality following the vaccination (p=0.28 comparing the pre-vaccination sleep quality to the 14-days and p=0.59 to the 3-month post-vaccination assessment, p=0.63 comparing both post-vaccination participations; Mann-Whitney U test, (*Figure 4A*). Similarly, in the **fourth vaccination** cohort, there was no change in sleep quality in the 14 days following the vaccination (p=0.56, Mann-Whitney U test, *Figure 4B*).

**Figure 4:**
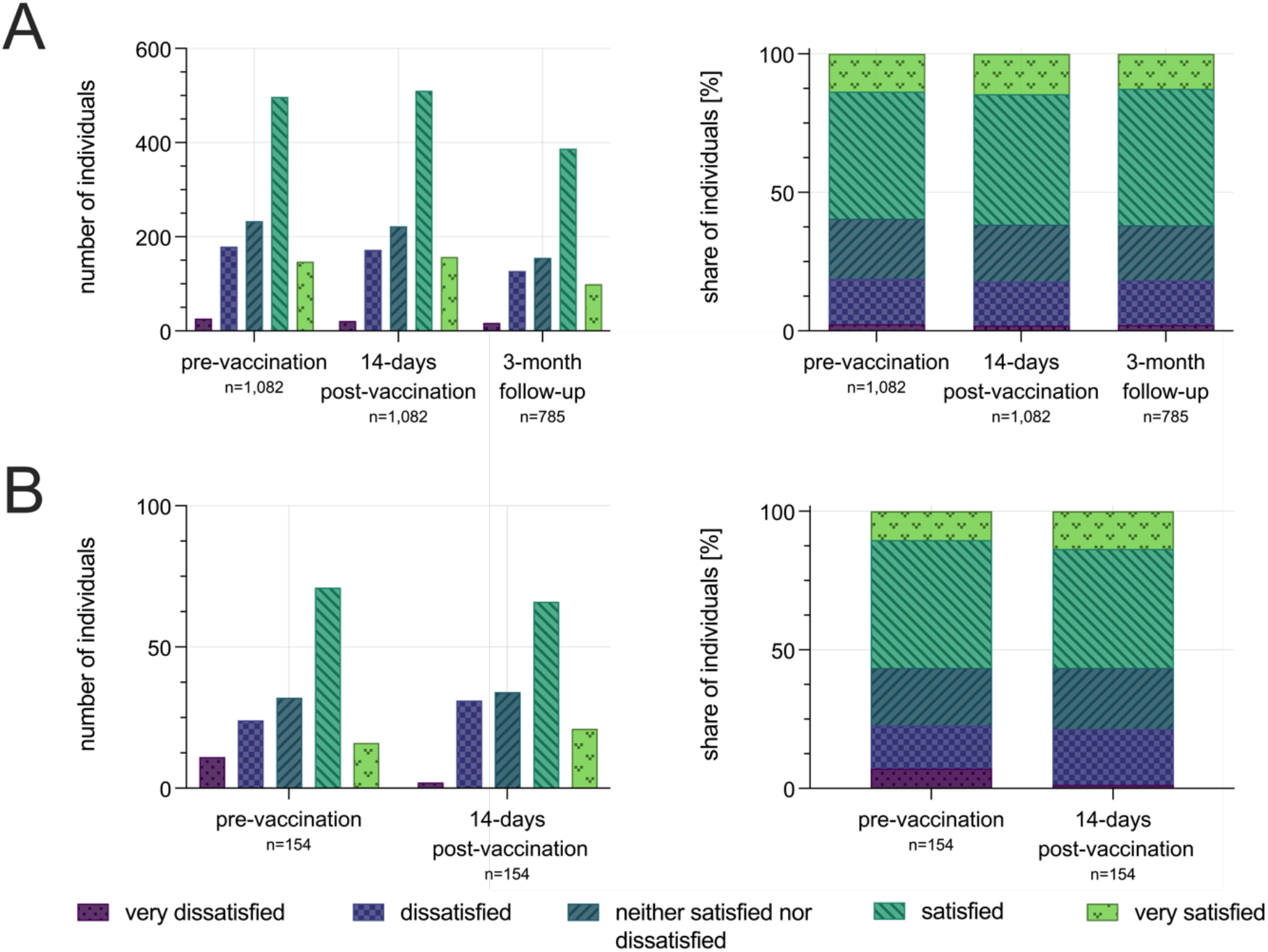
Sleep quality before and after COVID-19 booster vaccinations **A**) Sleep quality among HCWs before (pre-vaccination) and after (14-days and 3-month follow up) the third COVID-19 vaccination (left: absolute numbers, right: relative share). **B**) Sleep quality among HCWs before (pre-vaccination) and after (14-days) the fourth COVID-19 vaccination. Sleep quality is stratified in five categories (very dissatisfied to very satisfied).

## 4 Discussion

Mental health, sleep, and immune function are interlinked, and it is of great clinical relevance to clarify if individuals with psychiatric conditions or poor sleepers are at risk of an insufficient response to COVID-19 booster vaccinations or if booster vaccinations might deteriorate sleep quality. To our knowledge, this is the first study investigating the relationship between mental health, sleep, and immunogenicity of COVID-19 booster vaccinations. In our large sample of 1,082 individuals, no effect of psychiatric disorders or sleep quality on the immunogenicity of COVID-19 booster vaccinations and no effect of the booster vaccinations on sleep quality could be obtained.

Participants with psychiatric disorders had slightly but significantly lower antibody titres before the third COVID-19 booster vaccination compared to those without psychiatric disorders. However, HCWs with a psychiatric disorder were significantly older and had a higher BMI. Based on our regression analysis, the baseline differences in the Anti-SARS-CoV-2-Spike IgG levels can be explained by these two moderators and not by the presence of psychiatric disorders. At no other time point, Anti-SARS-CoV-2-Spike IgG levels differed between individuals with and without psychiatric conditions. This is an important finding, as it suggests that psychiatric conditions do not constitute a risk factor for an insufficient protection from COVID-19 following booster vaccinations.

Our null finding on the relationship between sleep quality and vaccine immunogenicity is in line with previous work that found an impact of sleep duration, but not of sleep quality or sleep efficiency on the post-vaccination humoral immune response for hepatitis B and influenza vaccinations (Prather et al., 2012; Prather et al., 2021) as well as with the finding that sleep disruptions resulting from sleep apnoea do not affect antibody titres following an influenza vaccination (Dopp et al., 2007). The difference between the clear effect of sleep restriction and sleep deprivation on vaccine immunogenicity (Dopp et al., 2007), at least in the short term (Benedict et al., 2012; Spiegel et al., 2002) and the absence of an impact of sleep quality or sleep efficacy might be explained by the large discrepancy between subjective and objective sleep variables, even in good sleepers (Benz et al., 2022).

Our null finding on the impact of the COVID-19 vaccinations on sleep quality provides another important result as it shows that in our large dataset there is no indication that COVID-19 vaccinations might be a precipitating factor for insomnia.

Our study has some important limitations. This large and representative example of HCWs represents a rather young and overall healthy population with only around 20% of participants reporting dissatisfaction with their sleep, while in the adult population dissatisfactory sleep is generally more common with more than 40% reporting some insomnia symptoms before the COVID-19 pandemic and more than 50% of the population during the pandemic in 2020 (Morin et al., 2021). The insomnia prevalence of 36% and the short sleep duration of only six hours in the first study investigating the association of sleep variables and the immunogenicity of COVID-19 vaccines might explain why the authors found a significant impact of the AIS and PSQI scores on antibody levels at some time points after adjustment for potential confounders (Athanasiou et al., 2023). It must be considered that both clinical scores include an item on sleep duration (Buysse et al., 1989). Another important limitation of our study is that the cohort is predominantly female. This is due to the recruitment of HCWs, which are mostly female in the German healthcare system (Gesundheitsberichterstattung des Bundes, 2021). It should further be mentioned as a limitation that for the fourth COVID-19 vaccination, data could only be collected for the time of analysis 14 days after the vaccine administration, since most follow-ups at 3 months post-vaccination and beyond have not yet been collected at the end of the data collection period for this manuscript in December 2022. The cohort for the fourth vaccination is also considerably smaller, as many individuals participating in the study for the third vaccination were not eligible for a second booster following the COVID-19 vaccination recommendations for HCWs of the German Standing Committee on Vaccination (STIKO)(Robert Koch-Institut (RKI), 2021). This is due to the fact that a relatively large proportion of the enrolled HCWs became infected with SARS-CoV-2 between the 3- and 6-month follow-up preventing an analysis of vaccine-induced increase of antibody titres. The majority of subjects received the third COVID-19 vaccination in autumn 2021. In the context of the high SARS-CoV-2 incidence phase, predominantly driven by the SARS-CoV-2 Omicron virus variant of concern in early 2022, a large proportion suffered a breakthrough infection (Bayerisches Landesamt für Gesundheit und Lebensmittelsicherheit, 2023). Regarding the study design, it should be highlighted that sleep quality was only recorded retrospectively for a 14-days time interval and based on the sleep quality item of the World Health Organisation Quality of Life (WHOQOL-BREF) questionnaire (Gholami et al., 2013) and not on a day-by-day self-report assessment with sleep diaries or an objective assessment with actigraphy or polysomnography. It should also be noted that due to data protection regulations, the study participants were not asked for a their specific psychiatric diagnosis and therefore it is not possible to investigate the impact of mood disorders on vaccine immunogenicity that has previously been found (Xiao et al., 2022).

Because of these limitations, mostly due to the real-life assessment in this large cohort study, our data does not exclude the possibility that severely disturbed sleep, insomnia, or specific mental health conditions might impact the immunogenicity of COVID-19 booster vaccinations. However, our results suggest that low sleep quality or mental health conditions in general have no relevant impact on antibody titres following COVID-19 booster vaccinations, especially compared to established mediators of COVID-19 vaccine immunogenicity, specifically age, smoking, and interval to the last immunising event (Reusch et al., 2022). While insomnia patients generally underestimate their sleep (Benz et al., 2022), polysomnographic measurements provide evidence for an objective reduction of sleep time in insomnia cohorts (Baglioni et al., 2014). Only studies including objective and subjective sleep measurements that are currently under way will be able to disentangle the impact of sleep quality and sleep duration on COVID-19 vaccinations (Lammers-van der Holst et al., 2022).

We interpret our data as a null finding suggesting that sleep quality and mental health status do not require adjustments of vaccination schemes. Expert advice had been given that shift workers should avoid vaccinations on shift days and hospital inpatients should consider rescheduling their vaccinations due to the sleep disruptions associated with hospitalization (Zhu et al., 2021). However, the first and only previous study investigating the effects of sleep on antibody levels following COVID-19 vaccinations found no correlation between the duration of sleep two days prior and one day after the vaccination with antibody levels (Athanasiou et al., 2023). In light of this and our data, and because COVID-19 vaccinations represent an essential disease prevention measure (Benenson et al., 2021), COVID-19 vaccines should be applied in a timely manner.

Given the relevant impact of sleep on immune function and the hesitancy of COVID-19 booster vaccination in large segments of the population, especially among psychiatric patients, our large cohort study provides important evidence for recommending COVID-19 booster vaccination in individuals with psychiatric disorders and sleep problems.

In conclusion, our large-scale real-world study on 1,082 individuals finds no effect of mental health and sleep quality on the immunogenicity of COVID-19 vaccinations and no change in sleep quality following vaccinations. Because this study was conducted in a rather healthy and young working population, it does not contradict previous experiments, demonstrating an impact of experimental sleep deprivation or sleep restriction, and a potential influence of insomnia disorder, on vaccination immunogenicity. However, our data suggests that sleep quality does not have a major impact on Anti-SARS-CoV-2-Spike IgG levels following the COVID-19 vaccination, especially compared to other established mediators of the vaccination response such as age, body weight and time to the last immunising event. Importantly, our data also indicates that COVID-19 vaccinations are not a precipitating factor for insomnia. While the role of sleep in the effectiveness of vaccinations should be investigated in greater detail, our findings can be interpreted as reassuring evidence against a major impact of mental health and sleep quality on the immunogenicity of COVID-19 vaccines or of the vaccine administration on sleep quality.

## Data Availability

Additional data that underlie the results reported in this article, after de-identification (text, tables, figures, and appendices) as well as the study protocol, statistical analysis plan, and analytic code is made available to researchers who provide a methodologically sound proposal to achieve aims in the approved proposal on request to the corresponding author.

## Acknowledgements

We thank the staff of the serological diagnostic laboratory for making their laboratory available and especially for their advisory help.

We explicitly thank Professor Ulrich Vogel, Infection Control and Antimicrobial Stewardship Unit, University Hospital Würzburg, Germany, for conception and design as well as funding support. He played a major role regarding the CoVacSer study but could not approve the final manuscript version as he died on the 4^th^ of October 2022. We miss him as an enthusiastic college and friend who showed a great dedication to his work, family, and friends.

## Funding

This study was funded by the Federal Ministry for Education and Science (BMBF) through a grant provided to the University Hospital of Würzburg by the Network University Medicine on COVID-19 (B-FAST, grant-No 01KX2021) as well as by the Free State of Bavaria with COVID-research funds provided to the University of Würzburg, Germany. Nils Petri is supported by the German Research Foundation (DFG) funded scholarship UNION CVD. Lukas B. Krone is supported by the Wellcome Trust (grant-No 203971/Z/16/Z) and Hertford College, Oxford, UK.

