## SupplementaryMaterial for "The relationship between mental health, sleep quality, and the immunogenicity of COVID-19 vaccinations"

### Supplementary Material

| Individuals threefold COVID-19 vaccinated | | | | | | | |
|---|---|---|---|---|---|---|---|
| | mentally healthy (n=921) | | underlying psychiatric disorder (n=161) | | total (N=1.082) | | p |
| | absolute | relative | absolute | relative | absolute number | relative number | |
| **How well can you enjoy your life?** | | | | | | | |
| *not at all* | 1 | 0.1% | 2 | 1.2% | 3 | 0.3% | <0.001 |
| *a little* | 10 | 1.1% | 11 | 6.8% | 21 | 1.9% | |
| *average* | 98 | 10.6% | 57 | 35.2% | 155 | 14.3% | |
| *quite* | 604 | 65.6% | 86 | 53.1% | 690 | 63.8% | |
| *extremely* | 208 | 22.6% | 5 | 3.1% | 213 | 19.7% | |
| **How well can you concentrate?** | | | | | | | |
| *not at all* | 0 | 0.0% | 2 | 1.2% | 2 | 0.2% | <0.001 |
| *a little* | 8 | 0.9% | 7 | 4.3% | 15 | 1.4% | |
| *average* | 156 | 16.9% | 59 | 36.6% | 215 | 19.9% | |
| *quite* | 584 | 63.4% | 90 | 55.9% | 674 | 62.3% | |
| *extremely* | 173 | 18.8% | 3 | 1.9% | 176 | 16.3% | |
| **Do you have enough energy for daily life?** | | | | | | | |
| *not at all* | 1 | 0.1% | 0 | 0.0% | 1 | 0.1% | <0.001 |
| *rather not* | 23 | 2.5% | 21 | 13.0% | 44 | 4.1% | |
| *halfway* | 118 | 12.8% | 57 | 35.4% | 175 | 16.2% | |
| *mostly* | 572 | 62.1% | 76 | 47.2% | 648 | 59.9% | |
| *completely* | 207 | 22.5% | 7 | 4.3% | 214 | 19.8% | |
| **Can you accept your appearance?** | | | | | | | |
| *not at all* | 1 | 0.1% | 5 | 3.1% | 6 | 0.6% | <0.001 |
| *rather not* | 16 | 1.7% | 20 | 12.4% | 36 | 3.3% | |
| *halfway* | 90 | 9.8% | 41 | 25.5% | 131 | 12.1% | |
| *mostly* | 543 | 59.0% | 75 | 46.6% | 618 | 57.1% | |
| *completely* | 271 | 29.4% | 20 | 12.4% | 291 | 26.9% | |
| **How satisfied are you with your ability to do everyday things?** | | | | | | | |
| *very dissatisfied* | 3 | 0.3% | 2 | 1.2% | 5 | 0.5% | <0.001 |
| *dissatisfied* | 23 | 2.5% | 21 | 13.0% | 44 | 4.1% | |
| *neither satisfied nor dissatisfied* | 87 | 9.4% | 41 | 25.5% | 128 | 11.8% | |
| *satisfied* | 526 | 57.1% | 82 | 50.9% | 608 | 56.2% | |
| *very satisfied* | 282 | 30.6% | 15 | 9.3% | 297 | 27.4% | |
| **How satisfied are you with yourself?** | | | | | | | |
| *very dissatisfied* | 0 | 0.0% | 5 | 3.1% | 5 | 0.5% | <0.001 |
| *dissatisfied* | 18 | 2.0% | 30 | 18.6% | 48 | 4.4% | |
| *neither satisfied nor dissatisfied* | 111 | 12.1% | 49 | 30.4% | 160 | 14.8% | |
| *satisfied* | 634 | 68.8% | 71 | 44.1% | 705 | 65.2% | |
| *very satisfied* | 158 | 17.2% | 6 | 3.7% | 164 | 15.2% | |
| **How satisfied are you with your sexual life?** | | | | | | | |
| *very dissatisfied* | 13 | 1.4% | 16 | 9.9% | 29 | 2.7% | <0.001 |
| *dissatisfied* | 44 | 4.8% | 18 | 11.2% | 62 | 5.7% | |
| *neither satisfied nor dissatisfied* | 175 | 19.0% | 48 | 29.8% | 223 | 20.6% | |
| *satisfied* | 474 | 51.5% | 64 | 39.8% | 538 | 49.7% | |
| *very satisfied* | 215 | 23.3% | 15 | 9.3% | 230 | 21.3% | |
| **How often do you have negative feelings such as sadness, despair, anxiety, or depression?** | | | | | | | |
| *never* | 199 | 21.6% | 4 | 2.5% | 203 | 18.8% | <0.001 |
| *not often* | 517 | 56.1% | 35 | 21.7% | 552 | 51.0% | |
| *occasionally* | 179 | 19.4% | 78 | 48.4% | 257 | 23.8% | |
| *often* | 26 | 2.8% | 41 | 25.5% | 67 | 6.2% | |
| *always* | 0 | 0.0% | 3 | 1.9% | 3 | 0.3% | |

**Supplementary Table 1**: Characterisation of mental health parameters separated by subgroups without and with psychiatric disorders undergoing a third COVID-19 vaccination

For each sub-cohort absolute numbers in the left. relative share in the right column. BMI: body mass index.

| Individuals fourfold COVID-19 vaccinated | | | | | | | |
|---|---|---|---|---|---|---|---|
| | mentally healthy (n=135) | | underlying psychiatric disorder (n=19) | | total (N=154) | | p |
| | absolute | relative | absolute | relative | absolute | relative | |
| **How well can you enjoy your life?** | | | | | | | |
| *not at all* | 0 | 0.0% | 0 | 0.0% | 0 | 0.0% | <0.001 |
| *a little* | 1 | 0.7% | 2 | 10.5% | 3 | 1.9% | |
| *average* | 18 | 13.3% | 8 | 42.1% | 26 | 16.9% | |
| *quite* | 83 | 61.5% | 9 | 47.4% | 92 | 59.7% | |
| *extremely* | 33 | 24.4% | 0 | 0.0% | 33 | 21.4% | |
| **How well can you concentrate?** | | | | | | | |
| *not at all* | 0 | 0.0% | 0 | 0.0% | 0 | 0.0% | <0.001 |
| *a little* | 2 | 1.5% | 2 | 10.5% | 4 | 2.6% | |
| *average* | 22 | 16.3% | 10 | 52.6% | 32 | 20.8% | |
| *quite* | 77 | 57.0% | 6 | 31.6% | 83 | 53.9% | |
| *extremely* | 34 | 25.2% | 1 | 5.3% | 35 | 22.7% | |
| **Do you have enough energy for daily life?** | | | | | | | |
| *not at all* | 0 | 0.0% | 0 | 0.0% | 0 | 0.0% | <0.001 |
| *rather not* | 4 | 3.0% | 2 | 10.5% | 6 | 3.9% | |
| *halfway* | 15 | 11.1% | 11 | 57.9% | 26 | 16.9% | |
| *mostly* | 85 | 63.0% | 6 | 31.6% | 91 | 59.1% | |
| *completely* | 31 | 23.0% | 0 | 0.0% | 31 | 20.1% | |
| **Can you accept your appearance?** | | | | | | | |
| *not at all* | 0 | 0.0% | 0 | 0.0% | 0 | 0.0% | <0.001 |
| *rather not* | 1 | 0.7% | 6 | 31.6% | 7 | 4.5% | |
| *halfway* | 17 | 12.6% | 3 | 15.8% | 20 | 13.0% | |
| *mostly* | 75 | 55.6% | 9 | 47.4% | 84 | 54.5% | |
| *completely* | 42 | 31.1% | 1 | 5.3% | 43 | 27.9% | |
| **How satisfied are you with your ability to do everyday things?** | | | | | | | |
| *very dissatisfied* | 0 | 0.0% | 1 | 5.3% | 1 | 0.6% | <0.001 |
| *dissatisfied* | 5 | 3.7% | 3 | 15.8% | 8 | 5.2% | |
| *neither satisfied nor dissatisfied* | 7 | 5.2% | 5 | 26.3% | 12 | 7.8% | |
| *satisfied* | 81 | 60.0% | 9 | 47.4% | 90 | 58.4% | |
| *very satisfied* | 42 | 31.1% | 1 | 5.3% | 43 | 27.9% | |
| **How satisfied are you with yourself?** | | | | | | | |
| *very dissatisfied* | 1 | 0.7% | 0 | 0.0% | 1 | 0.6% | <0.001 |
| *dissatisfied* | 5 | 3.7% | 3 | 15.8% | 8 | 5.2% | |
| *neither satisfied nor dissatisfied* | 14 | 10.4% | 8 | 42.1% | 22 | 14.3% | |
| *satisfied* | 87 | 64.4% | 8 | 42.1% | 95 | 61.7% | |
| *very satisfied* | 28 | 20.7% | 0 | 0.0% | 28 | 18.2% | |
| **How satisfied are you with your sexual life?** | | | | | | | |
| *very dissatisfied* | 1 | 0.7% | 0 | 0.0% | 1 | 0.6% | 0.0149 |
| *dissatisfied* | 7 | 5.2% | 1 | 5.3% | 8 | 5.2% | |
| *neither satisfied nor dissatisfied* | 32 | 23.7% | 10 | 52.6% | 42 | 27.3% | |
| *satisfied* | 64 | 47.4% | 7 | 36.8% | 71 | 46.1% | |
| *very satisfied* | 31 | 23.0% | 1 | 5.3% | 32 | 20.8% | |
| **How often do you have negative feelings such as sadness, despair, anxiety, or depression?** | | | | | | | |
| *never* | 37 | 27.4% | 0 | 0.0% | 37 | 24.0% | <0.001 |
| *not often* | 63 | 46.7% | 3 | 15.8% | 66 | 42.9% | |
| *occasionally* | 32 | 23.7% | 10 | 52.6% | 42 | 27.3% | |
| *often* | 3 | 2.2% | 5 | 26.3% | 8 | 5.2% | |
| *always* | 0 | 0.0% | 1 | 5.3% | 1 | 0.6% | |

**Supplementary Table 2**: Characterisation of mental health parameters separated by subgroups without and with psychiatric disorders undergoing a fourth COVID-19 vaccination

For each sub-cohort absolute numbers in the left. relative share in the right column. BMI: body mass index.
